# Comprehensive Evaluation of Artificial Intelligence Models for Diagnosis of Multiple Sclerosis Using Information from Retinal Layers Multicenter OCT Images

**DOI:** 10.1101/2024.03.05.24303789

**Authors:** Zahra Khodabandeh, Hossein Rabbani, Neda Shirani Bidabadi, Mehdi Bonyani, Rahele Kafieh

## Abstract

Multiple sclerosis (MS) is a chronic inflammatory disease that affects the central nervous system. Optical coherence tomography (OCT) is a retinal imaging technology with great promise as a possible MS biomarker. Unlike other ophthalmologic diseases, the variations in shape of raw cross-sectional OCTs in MS are subtle and not differentiable from healthy controls (HCs). More detailed information like thickness of particular layers of retinal tissues or surface of individual retinal boundaries are more appropriate discriminators for this purpose. Artificial Intelligence (AI) has demonstrated a robust performance in feature extraction and classification of retinal OCTs in different ophthalmologic diseases using OCTs. We explore a comprehensive range of AI models including (1) feature extraction with autoencoder (AE) and shallow networks for classification, (2) classification with deep networks designed from scratch, and (3) fine-tuning of pretrained networks (as a generic model of the visual world) for this specific application. We also investigate different input data including thickness and surfaces of different retinal layers to find the most representative data for discrimination of MS. Moreover, channel-wise combination and mosaicing of multiple inputs are examined to find the better merging model. To address interpretability requirement of AI models in clinical applications, the visualized contribution of each input data to the classification performance is shown using occlusion sensitivity and Grad-CAM approaches. The data used in this study includes 38 HC and 78 MS eyes from two independent public and local datasets. The effectiveness and generalizability of the classification methods are demonstrated by testing the network on these independent datasets. The most discriminative topology for classification, utilizing the proposed deep network designed from scratch, is determined when the inputs consist of a channel-wise combination of the thicknesses of the three layers of the retina, namely the retinal fiber layer (RNFL), ganglion cell and inner plexiform layer (GCIP), and inner nuclear layer (INL). This structure resulted in balanced-accuracy of 97.3, specificity of 97.3, recall 97.4%, and g-mean of 97.3% in discrimination of MS and HC OCTs.

## I. Introduction

Multiple sclerosis (MS) is a chronic inflammatory and neurodegenerative disease characterized by demyelination and neuro-axonal degeneration in the central nervous system (CNS) resulting in progressive tissue loss and worsening neurological disability over time[1]. Symptoms like fatigue, weakness, spasticity, vision changes, dizziness, mental changes, emotional changes, and depression are among the most prevalent in MS patients[2–4]. Various parts of the nervous system are affected in MS, including myelin sheaths, nerve fibers, and even cells that make myelin[5]. A few weeks are enough to recover if the injuries are not too severe, but severe injuries can permanently change the spinal cord[6]. Sclerosis refers to these permanent changes, which sometimes occur in multiple different parts of the body[7], thus giving rise to the name "multiple sclerosis". The cause of MS is unknown[8,9]. Scientists believe there is a combination of environmental and genetic factors involved in MS[10,11]. MS may be influenced by factors such as geography, vitamin D deficiency, obesity, and smoking[12,13]. The diagnosis of MS is established by applying the McDonald criteria[14], which primarily rely on the amalgamation of clinical, imaging, and laboratory evidence. However, it should be noted that the McDonald criteria are most applicable in cases where the disease has progressed to a relatively advanced stage. While magnetic resonance imaging (MRI) is the most established method to detect inflammations and lesions in MS[15], it is not sensitive or specific enough to reveal the degree of axonal damage[16].

MS causes extensive changes in the retina and optic nerve, which can be detected by a faster alternative method. Optical coherence tomography (OCT) is an imaging technology, which enables us to measure biomarkers such as thinning of individual layers in retina. [17,18]. The OCT technique uses in-vivo non-invasive imaging principles and is fast, with a temporal resolution of a few seconds. It allows for the precise delineation and measurement of various neuro-retinal layer thicknesses.

In particular, OCT has shown that the ganglion cell layer (GCL) has a gradual thinning from the early stages of MS, but the retinal nerve fiber layer (RNFL) serves as an indicator of axonal integrity[19–21]. As a potential biomarker for neurodegeneration, thinning of the peripapillary retinal nerve fiber layer (pRNFL), GCL, and inner plexiform layer thickness (IPL) are direct indicators of axonal damage[22,23]. Activation of inflammatory disease also alters the thickness of the inner nuclear layer (INL) [24]. These findings highlight the potential usefulness of measurements obtained from OCT as biomarkers for diagnostic applications in MS[25].

Artificial Intelligence (AI) has been widely used for automatic analysis of ophthalmic imaging modalities including OCT[26]. Instead of utilizing conventional statistical analysis, AI approaches can be used to predict the progression of MS-associated disability. Traditional methods for analyzing retinal layer thicknesses obtained through OCT entail extracting relevant features from the images and subsequently classifying them using specific techniques such as support vector machines (SVM)[27,28], linear discriminant function (LDF)[29], artificial neural network (ANN) [30–32], decision tree [32], logistic regression (LR) and logistic regression regularized with the elastic net penalty (LR-EN) [33], multiple linear regression (MLR), k-nearest neighbors (k-NN), Naïve Bayes (NB), ensemble classifier (EC), long short-term memory (LSTM) [34], and recurrent neural network [35].

Recent scientific investigations have explored the utilization of machine learning (ML) methodologies specifically focused on analyzing data pertaining to the thickness of the RNFL in each of the four quadrants delineating the peripapillary region (pRNFL), as well as GCL++ (between inner limiting membrane to INL), and the macular ganglion cell-inner plexiform layer (GCIPL). Furthermore, additional factors including age, sex, best-corrected visual acuity, and other relevant data were duly considered in the analysis[27–34]. In our prior research[36], we devised a machine learning approach to overcome limitations present in earlier automated methods for distinguishing between individuals with MS and HCs. To adhere to the principles of trustworthy AI, we improved interpretability by employing the occlusion sensitivity approach, allowing for the visualization of regional layer contributions to classification performance. The algorithm’s robustness was demonstrated through testing on a new independent dataset, confirming its effectiveness. Utilizing a dimension reduction method, we identified the most discriminative features from diverse topologies of multilayer segmented OCTs. Classification was carried out using SVM, Random Forest (RF), and ANN. The performance of the algorithm was evaluated through patient-wise cross-validation, ensuring that training and test folds encompassed records from distinct subjects[36].

Deep learning (DL) is a class of state-of-the-art ML techniques that has attracted a lot of global attention in recent years[37]. This simulates the operation of the human brain using numerous layers of artificial neural networks that can produce automatic predictions from input data. Unlike ML, the structure of DL uses more hidden layers to decode raw images without using manually creating specific features or feature selection algorithms. One of the notable attributes of deep learning networks is their capacity to deduce and extract latent and intrinsic features from data[26]. As a result, DL does not require any manual intervention at the stage of feature extraction, allowing the feature extraction and classification steps to be fully integrated into a Computer-Aided Diagnosis System (CADS)[38–40].

Convolutional neural networks (CNNs) are widely used in deep learning applications. They are inspired by the human visual system’s complex structure[41]. CNNs process images directly through convolutional layers, extracting features to create a predictive model. This eliminates the need for a separate detection model and provides an end-to-end solution that takes input OCT images and produces diagnostic decisions.

CNN systems have demonstrated strong diagnostic performance in applying ocular imaging, mostly OCT and fundus photographs[42]. Major ocular diseases which CNN methods have been applied for include diabetic retinopathy(DR) [43–45], glaucoma[46], age-related macular degeneration (AMD)[47,48], retinopathy of prematurity (ROP)[49], diabetic macular edema (DME), choroidal neovascularization (CNV), drusen, and others. Some of the classification methods based on CNN have been used to classify the OCT images in different ocular diseases[50–60]. Regarding MS, two studies have employed CNNs for classification. The first one that has utilized a CNN architecture for discriminating between individuals with MS and HC is [61], wherein CNNs feed with images of retinal layer thicknesses obtained with swept source OCT (SS-OCT) are used to diagnose early MS. To enhance the training dataset for CNNs, synthetic images representing retinal thickness are generated through the utilization of generative adversarial networks (GANs). This approach allows for the augmentation of the training data by producing synthetic images that capture variations in retinal thickness. The discriminative capability of the images is assessed through the utilization of effect sizes, specifically Cohen’s distance. Cohen’s parameter value is computed for each pixel in layers of retina. Pixels surpassing the predetermined threshold retain their thickness values, while those falling below the threshold are assigned a value of 0. Consequently, the input images for the CNN solely contain pixel information that is pertinent to the diagnosis of MS. As a result, the images fed into CNN only contain data on pixels that are pertinent to the diagnosis of MS.

In a recent investigation by Garcia Martin et al.[62], the Posterior Pole Protocol Spectralis SD-OCT, which incorporates an anatomic positioning system, was utilized for quantifying thickness measurements of the GCL, IPL, and RNFL. These measurements were subsequently employed as input data for a CNN designed for classification purposes. By using the mean thickness of the GCL (AUROC = 0.82) and the inter-eye difference in the IPL (AUROC = 0.71) as input features for a two-layer CNN, the automated diagnostic system achieved an overall accuracy of 0.87, a sensitivity of 0.82, and a specificity of 0.92.

In this study, we investigate an extensive array of AI models including (1) feature extraction with autoencoder (AE) and shallow networks for classification, (2) classification with deep networks designed from scratch, and (3) fine-tuning of pretrained network like ResNet152V2 for this specific application. In addition, we conduct an in-depth exploration of various input data, encompassing the thickness and surface measurements of different retinal layers, with the objective of identifying the most informative and representative data for discriminating MS. Moreover, we analyze the channel-wise combination and mosaicing techniques of multiple inputs, aiming to identify an optimal merging model that achieves improved performance.

This research explores the combination of different retinal layers, emphasizing the most effective composition of layers, specifically the mRNFL, GCIP, and INL. The results of this study are outlined in this manuscript. In order to meet the interpretability demands of AI models in clinical applications, we present the visualization of the contribution made by each input data to the classification performance. This is achieved through the utilization of occlusion sensitivity and Grad-CAM approaches. The data used in this study obtained from two independent public and local datasets. The efficacy and generalizability of the classification methods are substantiated through testing of the network on independent datasets. This evaluation showcases the ability of the methods to perform effectively beyond the initial training dataset. Our goal is to develop AI-based classification systems that can aid neurologists in the early detection of MS by analyzing alterations in sub-retinal layers of OCT.

## II. Materials and Methods

### A. Databases

The data used in this study includes 38 HC and 78 MS eyes that were achieved from two independent (but similar) datasets. The first dataset (Public dataset) has 14 HC and 18 MS and the second one (Local dataset) contains 24 HC and 60 MS. Demographic information of both datasets are included in Table 1. First, the models are trained and tested on the whole dataset and then, two datasets are used as separate training and testing datasets in measuring the robustness of the best-performing set of the classification algorithm.

**Table 1:** Demographic characteristics of participants of the Local and Public datasets.

| Dataset | Characteristics | HC | MS |
| --- | --- | --- | --- |
| Public | Current age, Y, mean ±SD | 35.77±13.03 | 41.97±8.77 |
|  | Sex, F, n(%) | 12(86%) | 14(78%) |
| Local | Current age, Y, mean ±SD | 26.3±3.06 | 34.5±8.03 |
|  | Sex, F, n(%) | 12(66%) | 30(85%) |

y: year, SD: standard deviation, n: number

#### Public dataset

The first dataset consists of 32 human retina scans taken with Spectralis SD-OCT equipment (Heidelberg Engineering, Heidelberg, Germany), 14 HC, and 18 MS OCTs which were manually segmented to eight retinal layers using internally developed software and reviewed and revised independently. At least, 12 images at the same location have been averaged for each Bscans, and the signal-to-noise ratio of the final averaged scans is at least 20 dB[63].

#### Local dataset

The second OCT dataset was collected between April 2017 and March 2019 at the Kashani Comprehensive MS Center in Isfahan, Iran[64]. The images were acquired using Spectralis SD-OCT and Heidelberg HEYEX version 5.1 by one trained technician with automatic real-time (ART) of 9 frames function for image averaging. The dataset consists of 24 HC and 60 MS eyes. All scans were checked for sufficient quality using OSCAR-IB criteria[65]. OSCAR-IB criterion is a standard for evaluating the quality of OCT images. Several indicators are considered as a quality indicator, forming the abbreviation OSCAR-IB: (O) obvious problems, (S) poor signal strength, (C) centration of scan, (A) algorithm failure, (R) unrelated retinal pathology, (I) illumination and (B) beam placement [65]. This criterion has been validated for MS [66].

### B. Data Preparation

Cross-sectional OCT scans have been effectively utilized in AI applications for the detection of various ophthalmologic diseases. However, the changes in sub-retinal thicknesses associated with MS are notably subtle, making it challenging to diagnose using unprocessed cross-sectional OCT scans alone. Hence, multilayer segmented OCTs are used rather than raw B-scans. Segmentation of the retinal layers is the key first step for AI-based analysis of MS by OCT images. Achieving precise and automated segmentation of retinal boundaries is crucial in this context, despite the notable challenges frequently encountered in images obtained from individuals with neurodegenerative diseases (NDD). These challenges include the presence of atrophic layers and ambiguous boundaries, media opacity, and artifacts resulting from patient movement. Overcoming these hurdles is essential for accurate analysis and diagnosis in such cases. The NDD-SEG presents a viable solution to this challenge. It is an optimal algorithm would possess the capability to effectively process various OCT devices and imaging protocols characterized by a wide range of resolution, size, artifacts, and noise[67]. So, the NDD-SEG approach is utilized for the segmentation of retinal layers in this study. By applying the NDD-SEG model, 9 boundaries and 8 layers of retina are extracted. The distances between pairs of retinal layers generates retinal thickness maps. After plotting the thickness maps, it becomes evident that certain maps exhibit destroyed regions. This destruction can be attributed to low quality, missed B-scan or the presence of noise in the B-scans used for generating the thickness maps. To address this issue and enhance the quality of the maps while simultaneously reducing noise, an inpainting algorithm is employed.

The data preparation steps are depicted in Figure 2 and are divided into the following sequential steps.

#### 1) Segmentation of retinal layers

The fundamental design of NDD-SEG[67] consists of a U-net structure that lacks pooling and upsampling layers. An initial network, referred to as the Retinal Tissue Segmentation Network (RTSN), proficiently identifies the presence of the retina within the image, even in the presence of a noisy background. Following this, the Retinal Layer Segmentation Network (RLSN) focuses on precisely segmenting the intra-retinal layers by utilizing the probability map derived from RTSN along with the original input image. In the decoder stages of RLSN, the probability map and texture features are integrated. Rather than a straightforward concatenation, the Self-Attention Transformer Block captures more advanced representations of the combined data. To maintain accurate layer boundaries, a novel Boundary Preservation Loss (BPL) function is introduced, allowing the incorporation of high-precision layer edges into a classification loss function. The integration of texture awareness and the focused preservation of edges imparts robustness to the network against noise. This approach achieves size-independence and resilient segmentation performance, even in the presence of image artifacts and pathologies. The Figure 1 presents a comparison between NDD-SEG and a conventional UNet approach for retinal layer segmentation in the presence of image artifacts and pathologies[67].

**Figure 1:**
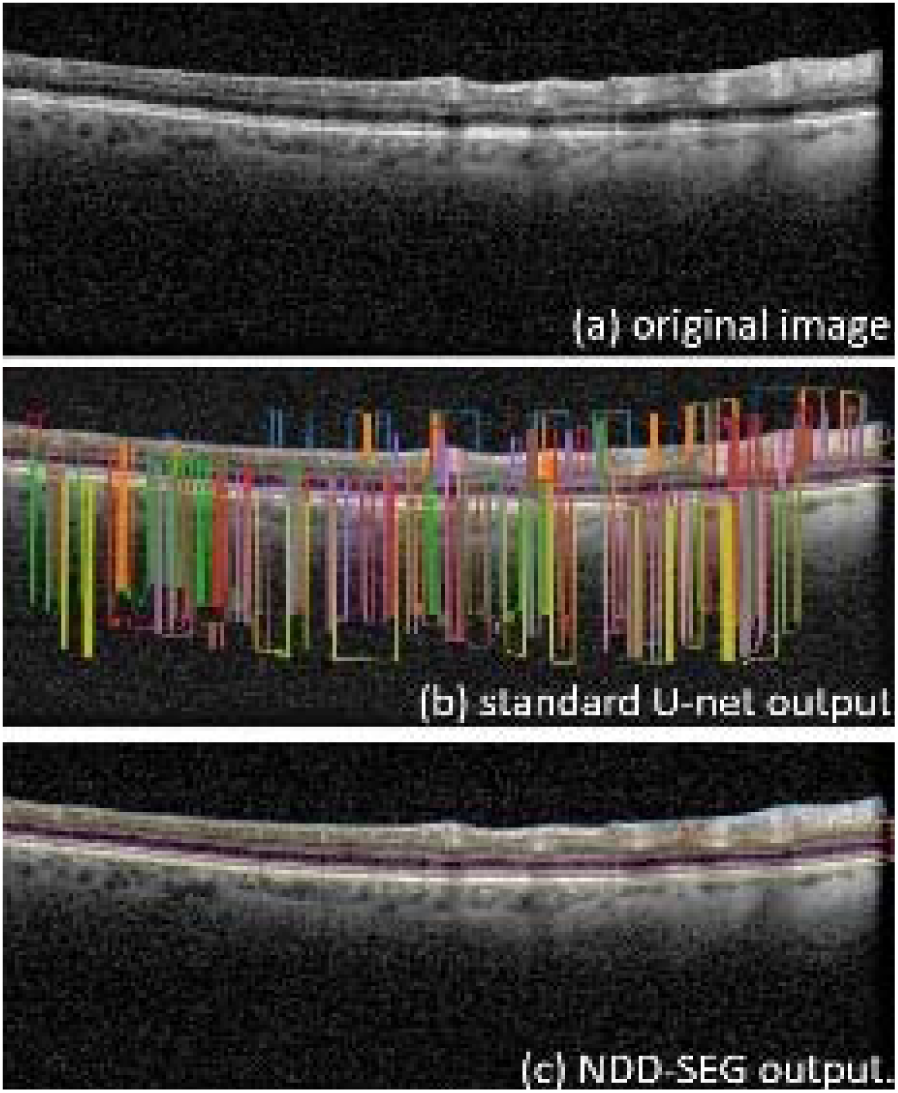
A comparison between NDD-SEG and a standard UNet approach for retinal layer segmentation [67]

#### 2) Extraction of thickness and surface of retinal layers

A noticeable change in the thickness of inner retinal layers, specifically the pRNFL and the combined macular ganglion cell and inner plexiform layers (mGCIPL), is evident when comparing individuals with MS to HCs[68]. Consequently, the assessment of inner retinal layer thicknesses serves as a diagnostic metric for distinguishing individuals with MS from HCs.

While changes in thickness can distinguish areas showing signs of retinal disease from normal regions, the use of texture descriptors can provide additional insights into disease progression. To identify neurodegenerative alterations using OCT image segmentation, the investigation focuses on variations in texture descriptors and optical characteristics of retinal tissue layers in patients with MS.[69]. Research indicates the potency of texture features in diagnosing MS cases[70]. Changes in retinal texture can result in modifications to the boundaries of the retina. As a result, measuring surface across different retinal layers offers an assessment method for differentiating individuals with MS from those unaffected.

#### 3) Quality Enhancement of the images

To enhance the quality of the images while simultaneously reducing noise, an inpainting algorithm is employed. Inpainting approach is used for quality enhancement and denoising in the data preparation step. In the inpainting algorithm, a highpass filter is initially applied to the input image, followed by the application of thresholding to generate a mask. Subsequently, Exemplar-based inpainting is employed to fill in the missing or corrupted regions of the image based on the mask.

The exemplar-based image inpainting approach utilizes a block matching algorithm to identify the most appropriate patch within the image that matches the target region. Once the optimal matching patch is identified, the pixel values from the source region are copied and pasted into the target region, effectively filling in the missing or omitted areas. The quality of the inpainted result is significantly influenced by the search for a matching patch in this technique. By selecting a patch that closely mirrors the content and structure of the missing region, the exemplar-based method aims to seamlessly integrate the inpainted region with the rest of the image[71].

#### 4) Feed images to network

In this study, we examine channel-wise combination and mosaicing of multiple inputs to find the better merging model. These combinations are shown in the final section of Figure 2.

**Figure 2:**
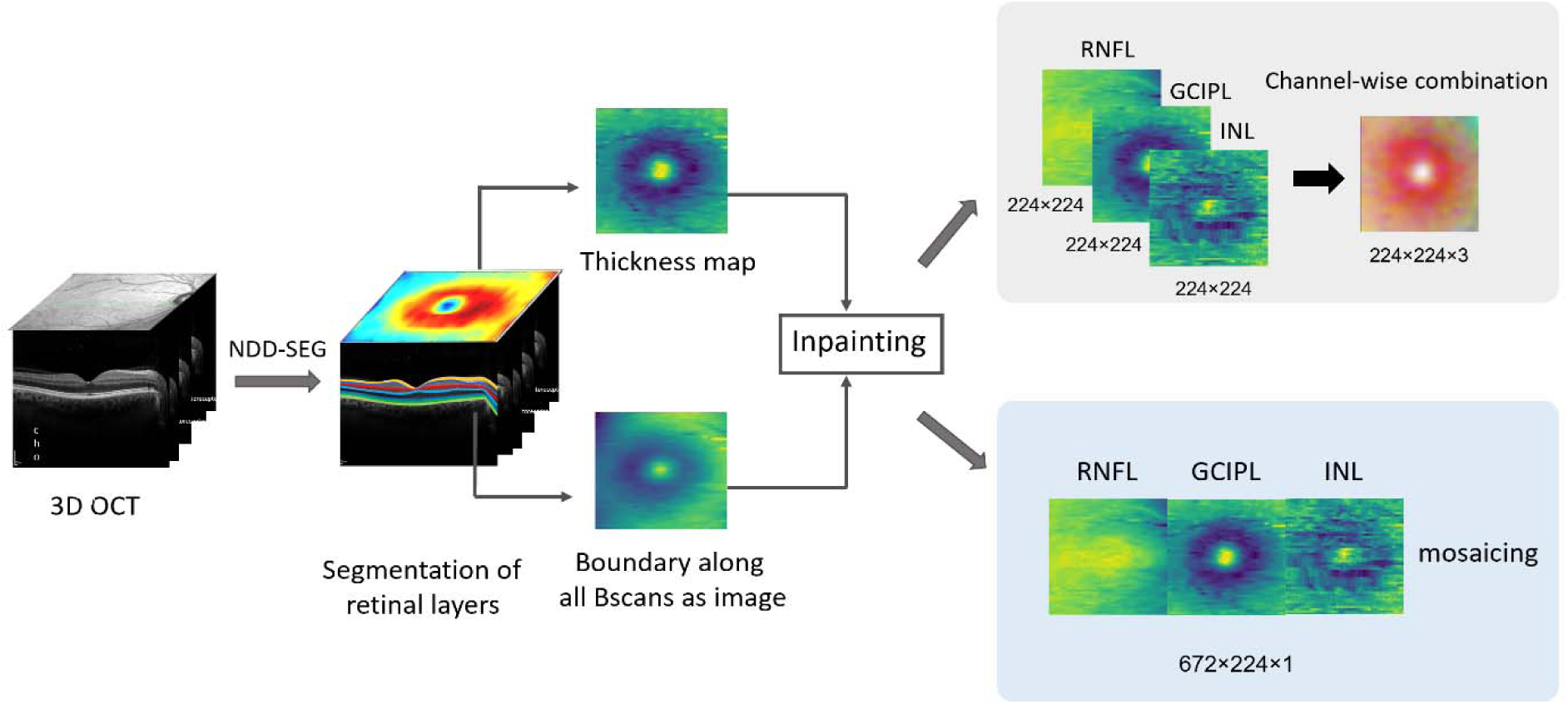
Input Data preparation steps

##### a) Channel-wise combination

To feed data into classifier models, we employ three thickness maps representing different retinal layers or three boundaries of retinal layers across all B-scans as individual channels in 3D color images. By exploring combinations of these retinal layers, our objective is to pinpoint the most effective discriminator for distinguishing between individuals with MS and HCs.

##### b) Mosaicing of multiple inputs

Another approach to identify the optimal merging model entails constructing a mosaic by combining three thickness maps or boundaries of retinal layers along all B-scans. Subsequently, a grayscale image is produced with the same width and three times the length, allowing additional analysis and comparison to determine the superior merging model.

### C. Classification methodology

In this research, we investigate a comprehensive range of AI models including (1) feature extraction using AE and shallow network for classification, (2) classification employing deep networks designed from scratch, and (3) fine-tuning of pretrained networks for classification.

#### 1) feature extraction with autoencoder (AE) and shallow network for classification

The architecture of the AE is designed to identify a low-dimensional representation for input images. An AE integrates an encoder function, responsible for altering the representation of input data, with a decoder function, which transforms the new representation back to the original format. The encoder compresses high-dimensional images into lower-dimensional displays, commonly referred to as the latent space or bottleneck representation. The decoder returns the data to its original dimensions. The latent space contains sufficient information to represent the data correctly. AEs are trained to preserve as much information as possible when input is sent through the encoder and subsequently the decoder[72]. Following the training of the AE and the reduction of features to the latent space, a shallow neural network is employed to classify the low-dimensional latent space.

When developing an AE, there are multiple possibilities to explore. The depth of an AE, often termed as the number of hidden layers in both the encoder and decoder networks, plays a crucial role. A deeper AE has the potential to learn more intricate representations, but this also increases the model’s capacity and computational requirements. The optimal depth is determined by the specific task, the complexity of the data, or a combination of both factors. To achieve the right balance between model complexity and performance, various depths are examined. The compressed representation of the input data is expressed through the latent layer of an AE, and the dimensions of this representation are contingent on the number of nodes in the latent layer. To effectively capture the essential information in the data without losing crucial details, the latent layer must have the appropriate number of nodes, a decision often influenced by the data’s complexity and the desired level of compression. While having too many nodes can result in overfitting or ineffective representation, too few nodes can result in information loss. In our classification framework, exploring the number of nodes in the shallow network is an additional option to be considered.

When investigating these options, hyperparameters are tuned and the model is choseb using the grid search method[73]. This method enables the exploration of various options to identify the optimal configuration.

#### 2) classification with deep network designed from scratch

In this study, a CNN model is utilized to achieve effective performance in the classification of OCT images. The network incorporates convolutional layers with Rectified Linear Unit (ReLU) activation functions, pooling, batch normalization, dropout, and fully connected (FC) layers. The last FC layer utilizes a Softmax activation function. Filters with size of 5, 4, and 3 are applied in various layers throughout the network. The nested-cross validation method is applied to select features and tune hyperparameters in CNN through the grid search method. With this approach, the classification model is chosen based on the outer fold that gives the maximum inner-fold accuracy[74]. The Adam optimizer and binary cross-entropy loss function are employed during training. The network architecture is adjusted throughout the training process to optimize performance. Due to the imbalanced dataset where the number of MS patients is more than HCs, class weight is incorporated during the model training processes.

#### 3) fine-tuning of pretrained Resnet152v2 network

Transfer learning (TL) in the context of CNNs is based on the premise that knowledge can be transferred at the parametric level. In this approach, pre-trained CNN models exploit the parameters of the convolutional layers to address a new task within the medical domain. Specifically, in TL with CNN for medical image classification, the task of classifying medical images can be accomplished by utilizing the generic features learned from a source task of natural image classification, where labels are available in both domains. There exist two approaches in TL for leveraging CNN models: the feature extractor approach and the fine-tuning approach. In the feature extractor approach, the convolutional layers are frozen, meaning their parameters are not updated during the model fitting process. Conversely, in the fine-tuning approach, the parameters of the convolutional layers are updated and adjusted during the model fitting process to adapt to the new task[75].

Numerous studies have consistently indicated that deeper models, such as ResNet (Residual Network) [76], exhibit superior performance compared to shallower models like VGG[75]. In the present study, we specifically investigated the performance of VGG16 and ResNet152V2 models, both pre-trained on the ImageNet dataset, through a process known as fine-tuning. Notably, ResNet152V2 demonstrated the most exceptional performance among the models evaluated, and therefore, its results are reported herein. Throughout the training process, the network architecture, including the number of nodes in the dense layers, was fine-tuned in order to attain optimal performance.

Figure 3 illustrates the AI models examined in the current study, encompassing the three approaches elucidated above.

**Figure 3:**
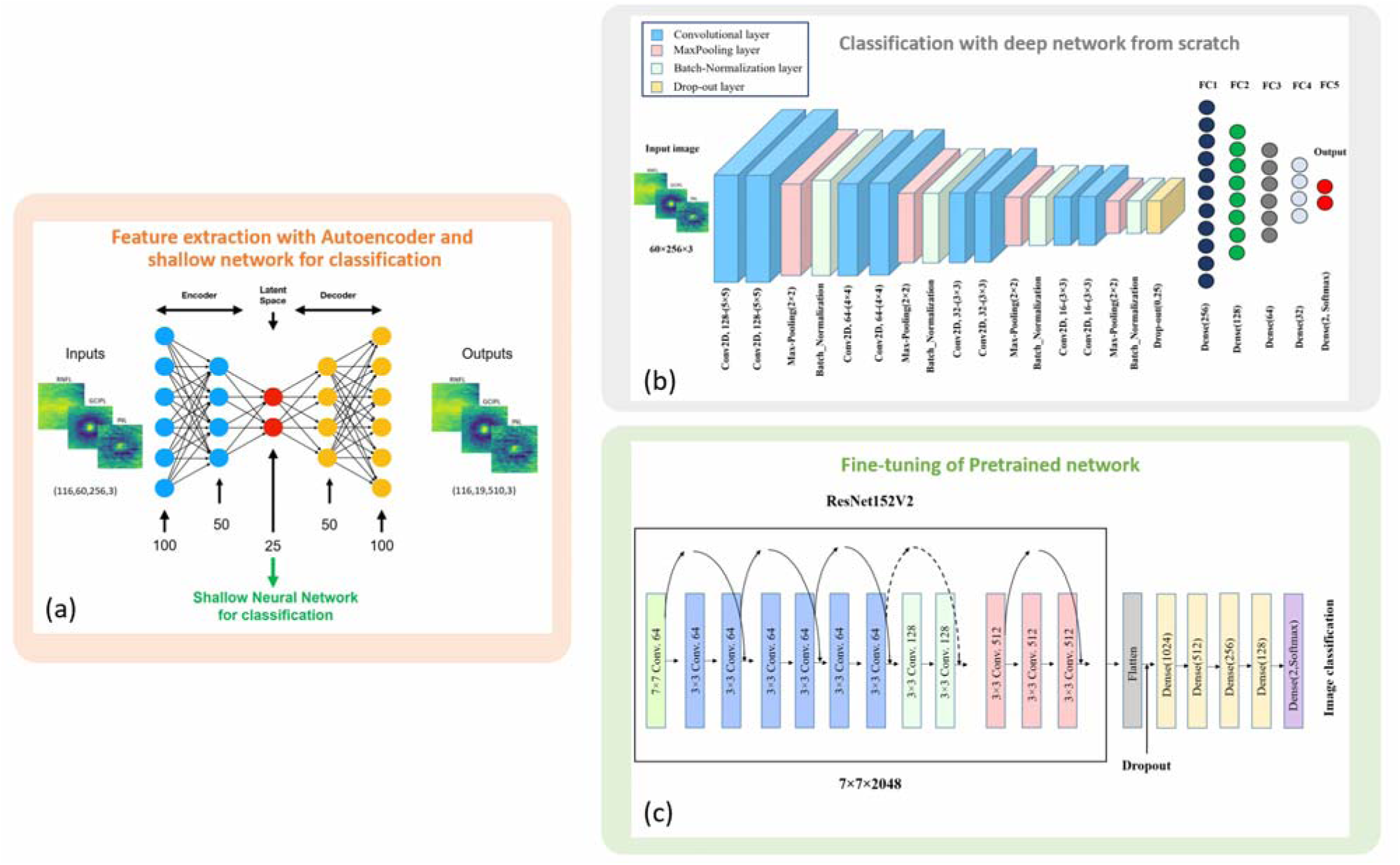
AI models for classification. (a) Feature extraction with autoencoder and shallow neural network for classification. (b) classification with deep network from scratch. (c) Fine-tuning of pretrained ReNet152v2 network

### D. Data Augmentation

#### 1) Data augmentation for the first model

After training the AE and reducing image features to the latent space, data augmentation is employed as a strategy to mitigate the risk of overfitting. Two widely utilized techniques for data augmentation, extrapolation and interpolation, are employed. Interpolation is the process of creating new data points within the range of previously collected data. It is frequently employed to generate more samples by interpolating between nearby data points. In contrast, extrapolation involves expanding the dataset beyond the current set of data points. This method is used to create samples outside the original data distribution by estimating or forecasting values based on patterns or trends that exist beyond the observable data. By combining interpolation and extrapolation techniques, data augmentation can effectively create new data instances with variations or characteristics similar to the original data.

#### 2) Image augmentation for the second and third models

Due to the limited size of our dataset, the likelihood of overfitting is heightened. To address this concern, Geometrical transforms such as horizontal and vertical flips, and randomly rotation with 20^0^, width shifting and height shifting with range of 0.1, zooming with range of 0.1 are implemented. These techniques aim to augment the dataset, thereby reducing the risk of overfitting.

## III. Experimental results

### A. Performance measures

Cross-validation, specifically employing a five-fold approach, serves as a means to assess results and prevent overfitting. This methodology guarantees that the ultimate outcomes remain unaffected by the initial data split. Within each fold, the training set undergoes augmentation, and the model is trained with these augmented sets, while the remaining set is reserved for testing the model without any augmentation. This process is repeated five times, altering the test set on each iteration. The final accuracy is determined by calculating the mean accuracy across these five distinct test sets.

In classification, the most widely adopted metric is accuracy, representing the rate of correct classifications. Nonetheless, there are instances, such as dealing with imbalanced datasets, where accuracy may not be the optimal choice. To address this, additional metrics such as G-means, F-measure, recall, and specificity are employed alongside accuracy to effectively handle imbalanced classifications[77].

The assessment of classification performance relies on the reporting of recall, g-mean, and specificity values, serving as quantifiable measures of the predictive capabilities of each model. The metrics are defined as follows:

Specificity: measures the ratio of negatives that are recognized correctly.

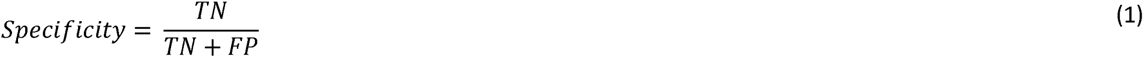

Recall: measures the positives ratio that are correctly recognized.

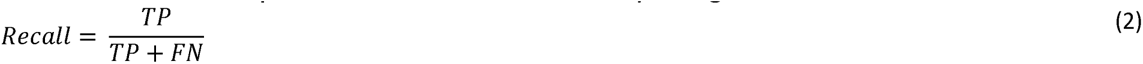

The quantities of true positives, false positives, false negatives, and true negatives are denoted as TP, FP, FN, and TN, respectively, with TP + TN + FP + FN = n representing the overall number of observations.

G-means: following recommendations from [78], the geometric mean (G-mean) is determined as the product of prediction accuracies for both classes, namely specificity (accuracy on the negative samples), and sensitivity (accuracy on the positive samples). A low G-mean value may be attributed to inaccurate categorization of the positive samples, even if the negative samples are classified correctly[79].

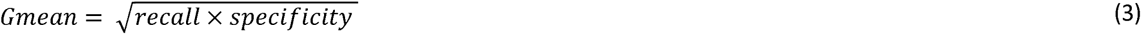

### B. Validation strategy

The nested cross-validation (nCV) technique begins with an initial division of data into outer folds, encompassing training and test categories. Following this, each outer training category undergoes further division into inner categories dedicated to setting hyperparameters (highlighted by the red box in Figure 3). Employing data augmentation and grid search techniques, the optimal internal training model is identified and subsequently applied to the external test data. The external model parameters, demonstrating a robust correlation with the highest accuracy achieved by the internal model parameters, are then chosen as the best model parameters[74] (Figure 4).

**Figure 4:**
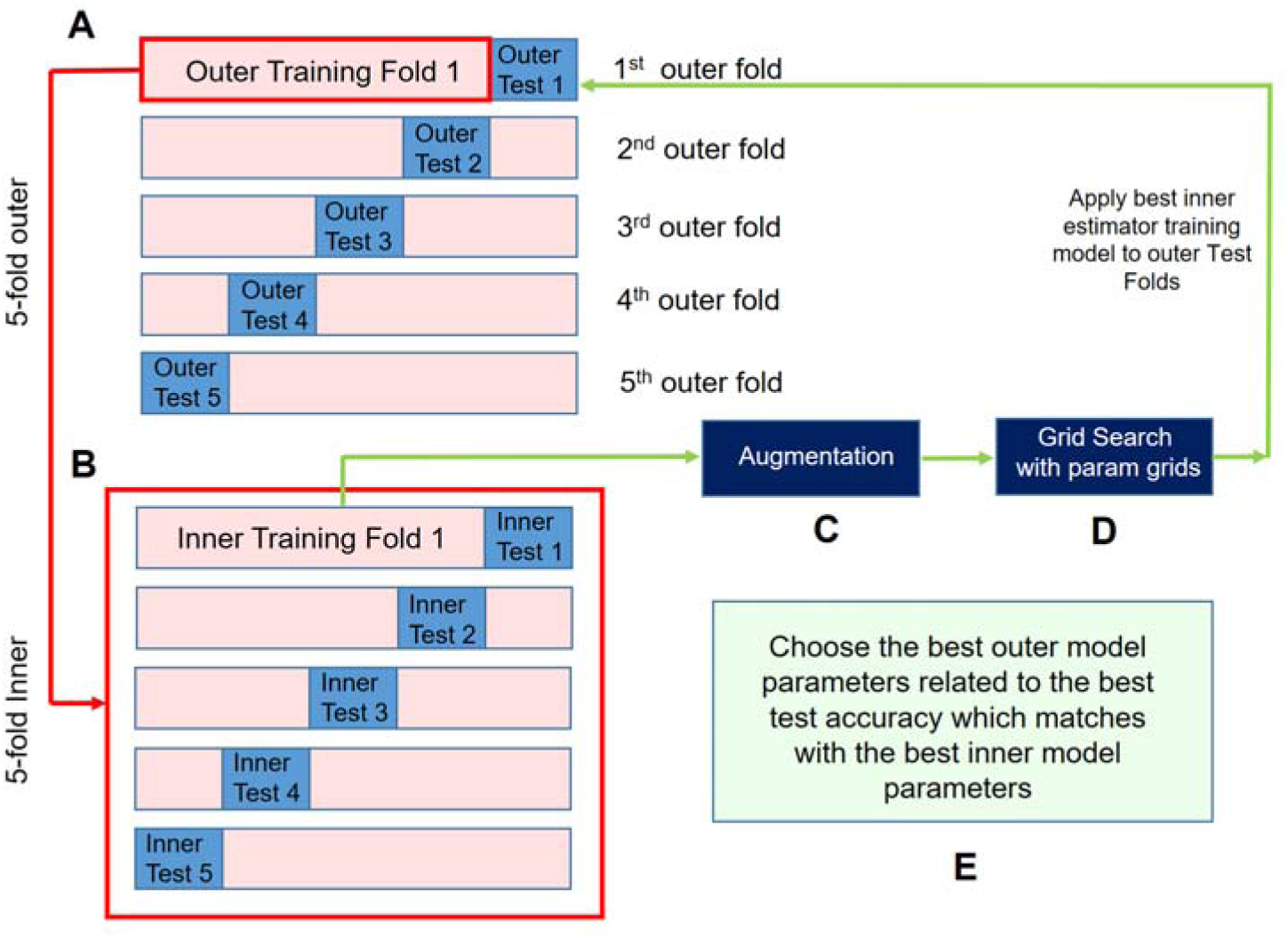
Nested-cross validation. (A) The data splits into outer folds containing train and test data pairs. (B) Outer training fold splits into inner folds. (C) Inner training fold augments to avoid overfitting. (D) Using grid search method for hyperparameter tuning. (E) Choose the best outer model matches with the best inner model parameters.

### C. Interpretability

Despite deep learning showcasing remarkable effectiveness across diverse tasks, particularly in image classification, interpretability has consistently presented a challenge for deep neural networks[80]. In this investigation, we enhance interpretability in deep learning algorithms through the classification process involving both a deep network built from scratch and fine-tuning of a pre-trained network. To address the inherent black box nature limitation of deep neural networks in medical applications, we employ occlusion sensitivity[81] and Gradient-weighted Class Activation Mapping (Grad-CAM)[82] methods.

Both occlusion sensitivity and Grad-CAM enhance the interpretability and clarity of deep learning networks. Occlusion sensitivity involves systematically blocking portions of the image to discern the significance of features, while Grad-CAM visualizes the areas of the image that wield the greatest influence in the model’s decision-making. These methodologies offer valuable insights into comprehending the internal mechanisms of deep learning models, aiding in the identification of discriminative regions or features crucial for predictions.

#### 1) Occlusion sensitivity

In this methodology, post-model training, a black mask measuring 40×40 pixels is generated and systematically applied to images within the test set, effectively traversing the entire image. The masked images are then fed into the model, and the resulting test accuracy is computed. A decrease in accuracy is expected if occlusion excludes regions containing crucial discriminative information. Reconstructing the occlusion with the original image size and accuracy values assigned to each pixel’s location illustrates interpretability, referred to as the heatmap. An interpretability heatmap delineates the localized impact on classification, highlighting the significance of each area in relation to the respective class.

#### 2) Grad-CAM

Grad-CAM generates a rudimentary localization map that accentuates specific areas in an image for concept prediction. This is achieved by leveraging the gradients of a chosen target concept as they flow into the concluding convolutional layer[82]. Utilizing the gradient information supplied to the final convolutional layer of the CNN, Grad-CAM discerns the significance of each neuron in relation to a specific decision[83].

### D. Performance results

The investigation involved the combination of various retinal layers through the implemented algorithms. In this section, the presented results encompass color images with resolutions of 60×256×3 and 224×224×3 pixels, where the thickness maps of the mRNFL, GCIP, and INL layers are allocated to three respective channels. Additionally, color images with resolutions of 60×256×3 and 224×224×3 pixels, incorporating the placement of the first three boundaries into their corresponding channels, along with grayscale images featuring a resolution of 60×768×1 pixels obtained by horizontally concatenating thickness maps, and grayscale images with a resolution of 60×768×1 pixels acquired through the horizontal concatenation of boundaries, are showcased. The outcomes of all combinations are detailed in Table 3. Initially, classification models were trained and tested on the complete dataset, comprising 116 OCT images, 38 HC images, and 78 MS images, utilizing 5-fold cross-validation.

**Table 3:** comparison of different models and structures for classification of MS and HCs.

|  | Channel-wise/<br>Mosaic | size | Data type<br>(Boundary/thickness) | Balanced-accuracy | Sensitivity | Specificity | g-mean |
| --- | --- | --- | --- | --- | --- | --- | --- |
| AE | Channel-wise | 60×256×3 | Thickness | 88.2% | 92.3% | 84.2% | 88.1% |
|  | Channel-wise | 224×224×3 | Thickness | 82.9% | 94.8% | 71.0% | 82.1% |
|  | Channel-wise | 60×256×3 | Boundary | 78.3% | 83.6% | 63.1% | 76.8% |
|  | Mosaic | 60×768×1 | Thickness | 84.9% | 88.7% | 73.6% | 84.1% |
|  | Mosaic | 60×768×1 | Boundary | 79.6% | 84.4% | 65.7% | 78.4% |
| From<br>Scratch | Channel-wise | 60×256×3 | Thickness | **97.3%** | 97.4% | **97.3%** | **97.3%** |
|  | Channel-wise | 224×224×3 | Thickness | 94.1% | 98.7% | 92.1% | 94.1% |
|  | Channel-wise | 60×256×3 | Boundary | 88.8% | 98.7% | 78.9% | 88.2% |
|  | Mosaic | 60×768×1 | Thickness | 93.4% | 97.4% | 89.4% | 93.3% |
|  | Mosaic | 60×768×1 | Boundary | 94.7% | 95.6% | 92.1% | 94.7% |
| Fine-tune | Channel-wise | 224×224×3 | Thickness | 94.0% | 96.1% | 89.4% | 93.9% |
|  | Channel-wise | 224×224×3 | Boundary | 91.5% | 92.2% | 89.4% | 91.5% |

To evaluate the models’ generalization capabilities, the classifiers that underwent training the public dataset, consisting of 14 HC and 18 MS samples, were subjected to testing on the local dataset, which includes 24 HC and 60 MS samples. Moreover, the local dataset was utilized for training purposes, while the public dataset was employed for testing. The corresponding outcomes are detailed in Table 4.

**Table 4.** generalizability of the methods by training the on the public dataset and testing on the local dataset and vice versa.

|  | Train data | Test data | Balanced-accuracy | Sensitivity | Specificity | g-mean |
| --- | --- | --- | --- | --- | --- | --- |
| AE | Public | Local | 84.5% | 83.3% | 85.7% | 84.5% |
|  | Local | Public | 79.3% | 94.4% | 64.2% | 77.9% |
| From scratch | Public | Local | 78.9% | 83.3% | 85.7% | 78.6% |
|  | Local | Public | 80.5% | 78.1% | 100% | 78.1% |
| Fine-tune | Public | Local | 66.2% | 70.0% | 62.5% | 66.1% |
|  | Local | Public | 66.6% | 83.3% | 50% | 64.5% |

#### Visual interpretability

The visual interpretability is depicted through the presentation of occlusion sensitivity and Grad-CAM heatmaps, as shown in Figure 5. To calculate the overlap of the heatmap and thickness maps, the formula heatmap × α + thickness map is utilized, with α set to 0.2.

**Figure 5:**
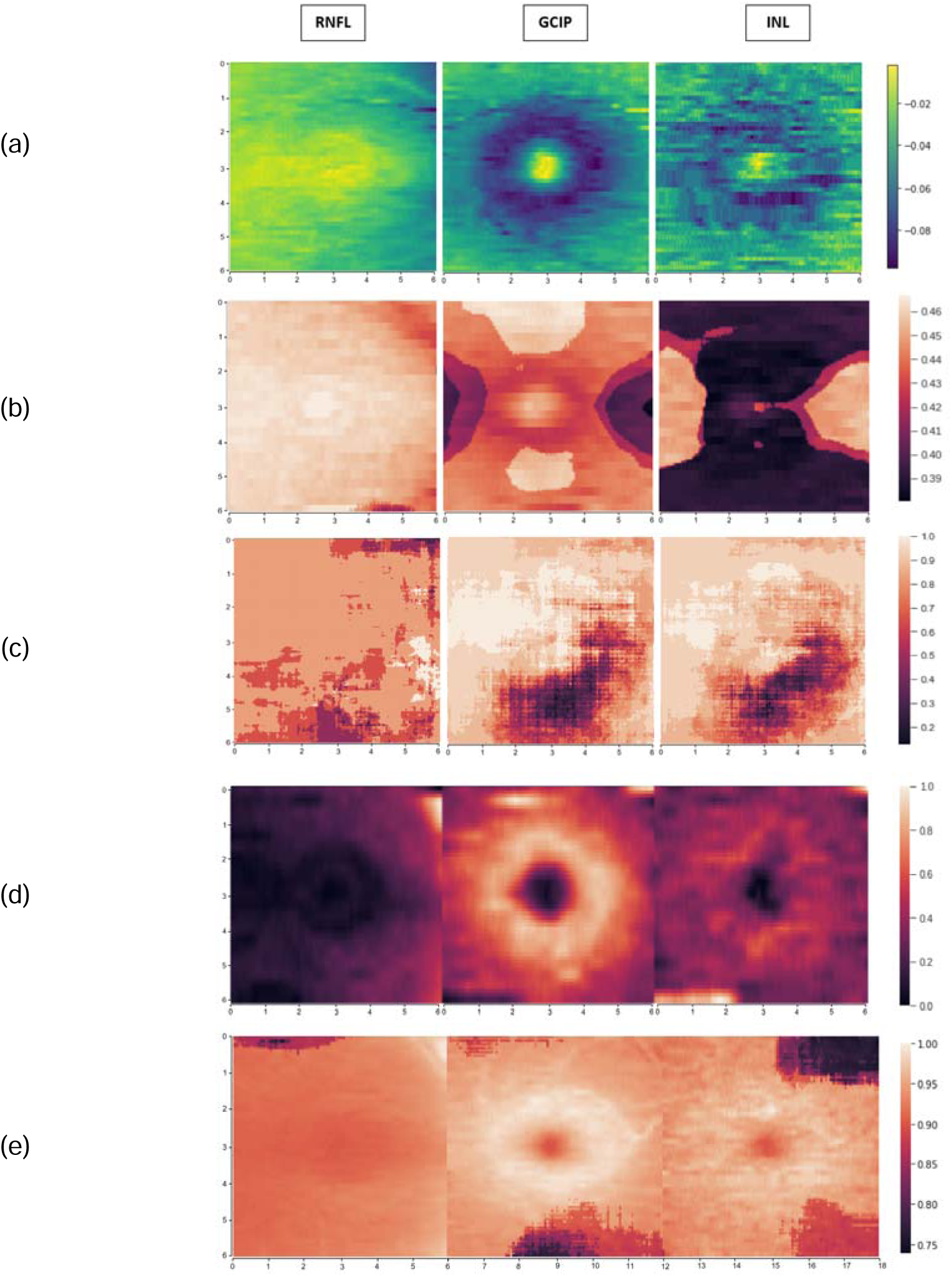
Visual Interpretability on Thickness Maps of mRNFL, GCIP, and INL Using Occlusion Sensitivity and Grad-CAM Methods. (a) Thickness maps in a sample from the MS dataset (x and y axes in millimeters). (b) Occlusion sensitivity heat map in MS and HC classification with the proposed CNN model trained from scratch and channel-wise combination of thickness maps. (c) Occlusion sensitivity heat map using the ResNet152V2 model and channel-wise combination of thickness maps. (d) Grad-CAM heat map with the proposed CNN model trained from scratch and mosaic of thickness maps. (e) Occlusion sensitivity heat map with the proposed CNN model trained from scratch and mosaic of thickness maps

For the proposed CNN network from scratch and fine-tuning with ResNet152V2, the visual interpretability is showcased through heatmap visualization methods, such as occlusion sensitivity and Grad-CAM. This is demonstrated in both channel-wise combination and the mosaic arrangement of images as input.

## IV. Discussion

In this work, we have explored a diverse array of AI models, encompassing (1) utilizing AE for feature extraction and shallow network for classification, (2) developing deep network from scratch, and (3) fine-tuning of pre-trained network for the purpose of classifying retinal OCT scans for discrimination of MS.

We additionally explored various input data, encompassing the thickness and surface of diverse retinal layers, in order to identify the most representative information for distinguishing MS. Furthermore, we examined the combination of multiple inputs through channel-wise combinations and mosaicing to determine an improved merging model.

The optimal topology for classification, employing the suggested deep network developed from scratch, was identified when the inputs comprised a channel-wise combination of the thicknesses of the mRNFL, GCIP, and INL retinal layers.

The reason for this outcome can be expressed that by combining the channels of images, the interconnection between the channels is established within the network. Given that neural-cellular, the layers are not independent of each other, by merging the channels of images, we intend for the network to establish connections between the layers, resulting in a better outcome.

As regards the analysis, it was observed that the inner retinal layers (mRNFL, GCIP, and INL) in the macular region exhibited the most significant impact, thereby demonstrating the highest capability to differentiate between control subjects and patients.

Utilizing a size of 60×256×3 among the employed channel wise combination combination, the proposed deep network developed from scratch yielded a balanced accuracy of 97.3%, specificity of 97.35%, recall of 97.4%, and a g-mean of 97.3% in discriminating between MS and HC OCTs. The execution time of an experiment for this model using Google Colaboratory, was calculated as 0.008 ± 0.007 seconds. This value was obtained by running the experiment five times, calculating the mean and standard deviation.

To prevent overfitting, data augmentation was employed in all networks. Moreover, patient-wise cross-validation was implemented in the training and testing datasets to avoid the leakage of testing data information into the training dataset. The study evaluated the generalizability of the proposed CNN model by training it on a local dataset and subsequently testing it on an independent Public dataset obtained from a new device in another country, and vice versa. The interpretability of the models was demonstrated through two approaches, namely occlusion sensitivity and Grad-CAM, to illustrate the contribution of regional layers to the classification performance. The heat maps generated by GCIP and INL revealed that the temporal and central regions had a greater impact on the classification of MS and HCs, as depicted in Figure 5 that was also achieved using machine learning algorithms in our previous study[36]. Therefore, the information from GCIP and INL layers in segregating MS and HC data, consistent with prior studies [22,24], was observed in interpretable outcomes of deep learning-based networks as well. Moreover, the thickness of the mRNFL was found to have a lesser impact on distinguishing MS disease when compared to GCIP.

Grad-CAM and occlusion sensitivity methods are common techniques for interpreting deep learning models. Despite sharing the same goal, they serve different purposes. Grad-CAM identifies crucial areas in an image by computing gradients of the target class relative to the feature maps in the network. The resulting heat map illustrates which portion of the image significantly influences the model’s prediction. On the other hand, the occlusion method systematically obstructs various regions of an input image and observes the impact on the model’s output. By assessing the change in the model’s prediction when different parts of the image are occluded, this method offers insights into the significance of various image regions.

To address age as a potential confounding variable between the MS and control groups, stochastic age-matching was applied to the test dataset before each model run. For every MS eye, the control eye with the closest age was included in the age-matched test dataset. This age-matching process was iterated five times for each model, and the outcomes were consolidated. This approach led to a well-balanced test dataset with no noteworthy differences in age between the MS and control groups.

The findings indicate that each protocol has its own merits depending on the dataset size. Specifically, for smaller datasets, the combination of feature extraction using AE and a shallow neural network demonstrates superior performance. On the other hand, deep learning-based methods provide interpretability in the results.

Table 6 shows a summary of the previous studies on ML and DL algorithms in distinguishing MS and HCs.

**Table 6.** Summary of previous studies.

| Previous works | Number of datasets | Input retinal layers | Performance metrics | The most discriminant retinal layer | Classification method |
| --- | --- | --- | --- | --- | --- |
| Garcia-Martin et al. [29] 2012 | 115 MS, 115 HC | Peripapillary area | Specificity=95% | pRNFL | Linear Discriminant Function (LDF) |
| Garcia-Martin et al. [30] 2013 | 106 MS, 115 HC | Peripapillary area | AUC=0.945 | pRNFL | ANN |
| Garcia-Martin et al. [31] 2015 | 112 MS, 105 HC | Peripapillary Area | Recall=89.3%<br>Specificity=87.6% | pRNFL | ANN |
| Palomar et al. [32] 2019 | 80 MS, 180 HC | Peripapillary, macular and extended (between macula and papilla) areas | Decision tree in the macular area:<br>AUC=0.959<br>In the extended area:<br>AUC=0.998 | pRNFL | Decision tree, ANN, SVM |
| Cavaliere et al. | 48 MS, | Peripapillary and | Recall=89% | GCL++ | SVM |
| [27]<br>2019 | 48 HC | macular areas | Specificity=92% | and nasal quadrant<br>of the outer and<br>inner ring in pRNFL |  |
| Garcia-Martin<br>et al. [28]<br>2021 | 48 MS,<br>48 HC | Macular area | Recall=98%<br>Specificity=98% | GCL++ | SVM,<br>ANN |
| Zhang et al.<br>2020 | 58 MS,<br>63 HC | Macular area | Recall=64%<br>Specificity=94% | GCIPL | logistic regression<br>(LR), logistic<br>regression regularized<br>with the elastic net<br>penalty (LR-EN), SVM |
| Montolio et al.<br>[34]<br>2021 | 108 MS,<br>104 HC | Peripapillary and<br>macular areas | EC:<br>Recall=87%<br>Specificity=88.5%<br>LSTM:<br>Recall=81.1%<br>Specificity=82.2% | pRNFL | MLR), SVM, decision<br>tree, k-NN, NB, EC,<br>LSTM recurrent neural<br>network |
| López-Dorado<br>et al.<br>[61]<br>2022 | 48 MS,<br>48 HC | Macular area | Specificity = 100%<br>Recall=100% | GCL++, whole retina<br>GCL+ (Cohen<br>distance) | Augmentation with<br>GAN and classification<br>with CNN |
| Khodabandeh<br>et al. [36] 2023 | Train: 106 MS,422 HC<br>(Charite dataset)<br>Test: 67 MS, 45 HC<br>(Isfahan dataset) | Macular area | Precision = 89%<br>Recall =88% | GCIPL and INL | SVM, RF, and ANN |
| Garcia-Martin<br>et al. [84]<br>2023 | 100 MS,<br>111 HC | Posterior pole<br>grid 8×8 and zone<br>grouping | Recall = 90.90 %<br>Specificity = 95% | RNFL and GCL | Gradient boosting, RF,<br>Explainable boosting<br>machine |
| Garcia-Martin<br>et al. [85]<br>2023 | 79 MS,<br>69 HC | Inter-eye<br>difference using<br>posterior pole<br>protocol | Recall = 86%<br>Specificity = 90 % | RNFL, GCL, and IPL | SVM- RFE- Leave-One-<br>Out cross validation<br>(SVM-RFE-LOOCV) |

Previous studies lacked direct utilization of images as input for models, and there was a scarcity of deep learning methodologies employed in those investigations. The results of this study cannot be directly compared to previous works due to the unavailability of codes and datasets in those studies. It is important to note that this study did not consider the presence of optic neuritis (ON), and therefore, MS patients with and without ON were combined for classification. Consequently, the lower performance compared to studies considering MS with ON is reasonable, as eyes without ON exhibit less thinning and are less distinguishable from healthy controls. Finally, some previous studies incorporated pRNFL data as input for classification, resulting in higher performance compared to the limited focus on the macular region in this study.

This study has several limitations. Firstly, it did not consider the history of ON, a prevalent clinical symptom of MS. Secondly, longitudinal follow-up of patients was not conducted, which could aid in the early diagnosis of the disease. Thirdly, the models were trained using data from a specific device, and it was not evaluated if the models are generalizable to other devices, such as TOPCON or ZEISS. To improve the model’s ability to interact with new datasets, additional data from different devices should be included in the training dataset in future studies. We anticipate that addressing these limitations will lead to better results and a clearer demonstration of the generalizability of the proposed model.

## Data Availability

Two datasets used in this manuscript were obtained from the following two references:
local dataset:
https://www.sciencedirect.com/science/article/abs/pii/S2211034820306994
public dataset:
https://www.ncbi.nlm.nih.gov/pmc/articles/PMC6327073/

https://iacl.ece.jhu.edu/index.php?title=Main_Page

## Notes

### Competing Interest Statement

The authors have declared no competing interest.

### Author Declarations

The local dataset was approved by the regional bioethics committee in Isfahan University of Medical Sciences and written informed consents were obtained after participants were informed about the aims of the study.

